# Modeling the Heterogeneity and Trajectories of Cognitive Dysfunction in Parkinson’s Disease Using Partially Ordered Set Models

**DOI:** 10.1101/2025.07.21.25331873

**Authors:** Cole Zweber, Judith Jaeger, Cyrus Zabetian, Rebecca Miller, Vivikta Iyer, Amie Hiller, Satya Sahoo, Brenna Cholerton, Abigail Ryan, Curtis Tatsuoka, Deepak K. Gupta

## Abstract

Traditional binary classifications of Parkinson’s disease (PD) cognitive dysfunction fail to capture its inherent heterogeneity. This study introduces the Partially Ordered Set (POSET) model, a Bayesian framework, to analyze cognitive trajectories using Parkinson’s Progression Markers Initiative (PPMI) data. Five cognitive domains: Attention, Visuospatial Judgement, Executive Functioning, Working Memory, and Episodic Memory, were mapped onto nine neuropsychological measures to calculate Cognitive Performance Scores (CPS). Of 264 patients without baseline cognitive dysfunction, 21.7% developed dysfunction by Year 3. These individuals exhibited significantly lower median CPS across all domains during follow up visits in Years 1-3. Notably, baseline Attention and Visuospatial CPS were significant predictors of future impairment, with an area under the curve (AUC) of 0.782; a specificity of 91.3%, and a sensitivity or 35.7%. POSET modeling provides a sophisticated approach to characterizing PD cognitive decline, offering greater granularity than conventional schemes. Further large-cohort studies are needed to confirm these findings.

## Introduction

Cognitive dysfunction is one of the quintessential non-motor features of Parkinson’s disease (PD)^1^, occurring along a spectrum from mild cognitive impairment (PD-MCI)^2, 3^ to dementia (PD-D)^4, 5^. Using the conventional PD-MCI and PD-D frameworks^6^, cognitive dysfunction is estimated to be twice as common in PD patients compared with the general population^7^, affects 25-30% of newly diagnosed PD patients^8^ and impacts up to 80% of PD patients after 20 years of disease duration^9^. Incidence, prevalence, and severity of PD-MCI and PD-D increase with advancing age and disease duration, with age being the strongest risk factor^10^. Importantly, PD-MCI in the early stages of disease predicts later development of PD-D^11, 12^ and is associated with increased mortality^13^. Furthermore, cognitive dysfunction in PD is a highly burdensome non-motor symptom for patients and caregivers and remains challenging for clinicians with symptomatic medications and lack of disease modifying medications ^14^. As for the neurobiological basis of cognitive dysfunction in PD^15^, several risk factors or biomarkers have been associated with PD-MCI or PD-D, including male sex^16^, orthostatic hypotension, glucocerebrosidase (GBA) mutation^17^, triplications in the α-synuclein gene, and low amyloid-β42 in cerebrospinal fluid^18, 19^, although no marker has demonstrated sufficient accuracy to reliably predict progression to PD-MCI or PD-D.

Currently used schemes of PD-MCI and PD-D for investigating and classifying cognitive dysfunction in PD^20^ are inherently restricted because they provide limited analysis of the underlying neuropsychological data and rely on binary (yes/no) outcome measures. Consequently, such categorical frameworks fail to capture the heterogeneity and the continuum of cognitive dysfunction in PD. Accordingly, there is a critical need for approaches that treat cognition as a continuous, multidimensional construct, enabling more granular subtyping and characterization of trajectories. This is needed to better counsel patients and families on likely disease progression, and to improve the efficiency and accuracy of clinical trials.

In this paper, we will 1) review PD-MCI and PD-D classification schemes (including their limitations with respect to interpreting neuropsychological data) 2) review Partially Ordered Set (POSET) modeling and 3) apply POSET modeling to the Parkinson’s Progression Markers Initiative (PPMI) data (to illustrate a novel approach to characterizing cognitive dysfunction in PD).

Conceptually, Mild cognitive impairment (MCI) refers to a syndrome representing a stage between normal aging and dementia. Historically, MCI was defined as a transitional stage of cognitive decline, initially described in the context of Alzheimer’s disease and later extended to other etiologies, including PD. Dementia generally refers to an advanced stage of cognitive dysfunction when it becomes severe enough to negatively affect social and/or occupational functioning and thus cause impairment in functional independence^21^. Although the two classification schemes share several data elements for classifying cognitive dysfunction in PD, a patient can only be classified as either PD-MCI or PD-D (Table 1), provided there are no other causes of the impairment (e.g., delirium, medication effects, etc.).

**Table 1:**
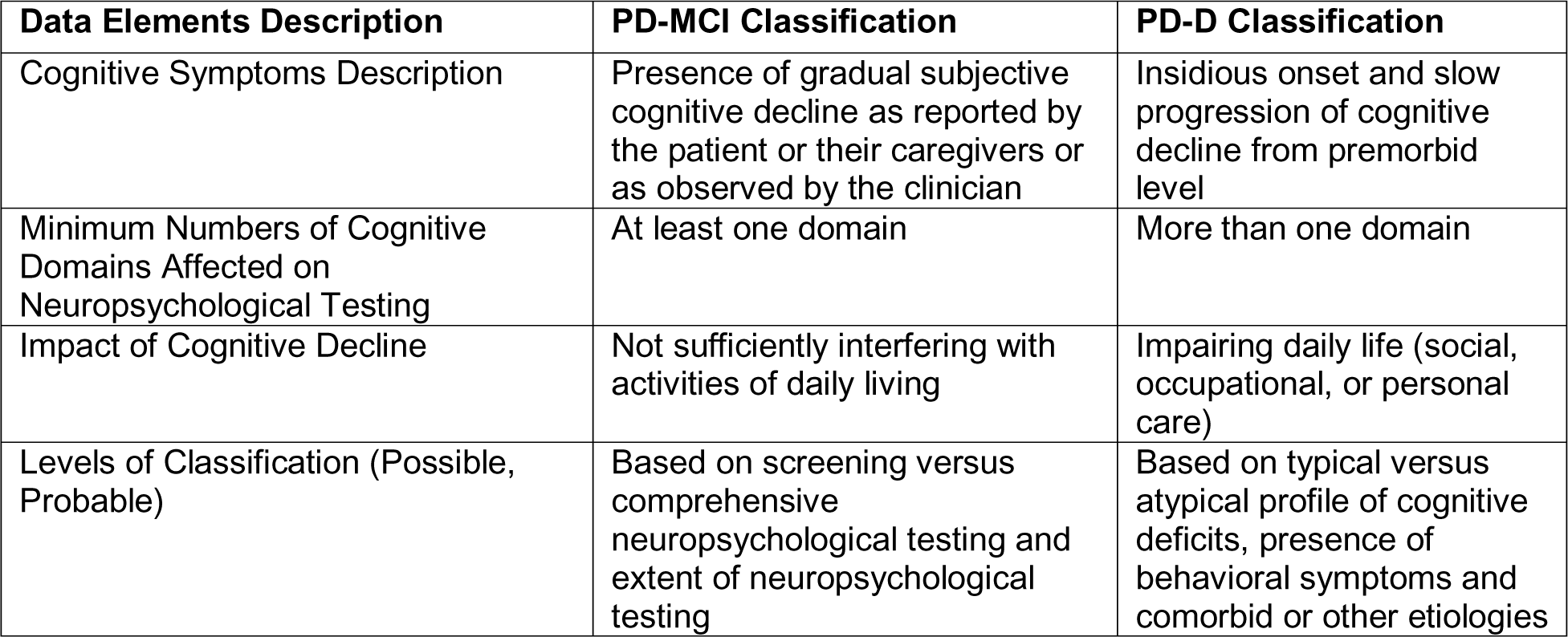
Comparing PD-MCI and PD-D Classification. Comparison of PD-MCI and PD-D classification schemes for describing cognitive dysfunction of a PD patient in mutually exclusive categories.

A cognitive domain is a broadly defined area of cognition (e.g., memory, executive functioning) while specific cognitive functions are more narrowly defined aspects of ability^22^. For example, within the domain of memory, evaluators assess multiple discrete functions including encoding, free recall, and recognition discrimination. Neuropsychological tests often engage multiple cognitive functions across cognitive domains. Although neuropsychological tests are categorized into domains based upon the primary function they are assessing, other cognitive functions influence overall test performance. The overlapping involvement of multiple cognitive domains and functions on any given neuropsychological measure is difficult to minimize and can make understanding the true meaning of test scores more challenging. Some neuropsychological tests, such as memory measures, produce multiple scores that reflect these different functions, but many tests only produce one total score. To complicate matters further, cognitive tests also tap into functions that may be classified under a different cognitive domain. Keeping with the example of memory testing, these measures also reflect an individual’s attention, executive functioning, and language or visuospatial abilities. In research, similar challenges can arise due to the necessity of strictly defined cognitive domains. In clinical practice, clinicians utilize more comprehensive test batteries to look for patterns within and across test domains to help clarify the meaning of test scores.

Interpretation and analysis of neuropsychological data using the PD-MCI and PD-D classification schemes present a complex problem when describing cognitive dysfunction. As noted above, a problem in linking performance on a test item to distinguish impairment in a certain cognitive domain is that most items often require engagement with more than one of the cognitive functions within that domain and may involve additional other functions to perform well. If a patient performs poorly on an item involving several cognitive functions, it can be unclear which one or more of the involved functions may be impaired. Despite the wide variety of cognitive batteries available, clarifying specific deficits in cognitive domains is complicated by the challenge of effectively delineating which cognitive functions are required to perform well on a test item mapped to that domain. Poor performance on a given task may stem from a variety of specific deficits; for instance, subpar performance on a verbal list-learning task may be attributed to deficits in verbal fluency, typically classified as an executive dysfunction, or due to impaired attention. These overlapping domains of cognitive dysfunction have been ineffectively addressed using subscales—an attempt to increase specificity in attribution of impaired functions to cognitive performance by ignoring the multi-faceted nature of tasks. Subscales rely on simplistic assumptions that there are one-to-one correspondences between a task and a specific function, which may obfuscate the true cognitive underpinnings of poor performance. In addition, reliance on total scores from expanded cognitive batteries presents similar issues. While the summed scores may indicate the presence or absence of cognitive deficits relative to norms, it is possible to produce the same summed score from multiple iterations of responses and/or task performances, further obscuring the involvement or absence of specific deficits.

POSET models^23^ permit classification of the participants’ functioning with respect to specific cognitive functions^24, 25^ and have been applied to predict conversion from MCI to Alzheimer’s disease^26–28^ and distinguishing neurocognitive profiles in schizophrenia^29^. POSET modeling addresses aforementioned limitations of current approaches to neuropsychological scoring. Classification is conducted based on observed responses to neuropsychological measures after each measure is mapped to cognitive operations that are involved in performing well on that measure, thereby making the link between observed responses and latent functioning explicit. This key step is conducted data- analytically through assessing consistency of specifications to response data^30^, with additional input from three expert neuropsychologists. A partially ordered set of possible profiles of performance (discrete levels, such as high or low) across cognitive functions can be generated. Partial orders provide flexible representation of response patterns, allowing individuals to exhibit distinct profiles of strengths and weaknesses rather than being constrained to a single linear severity continuum. Because each cognitive battery invokes multiple specific cognitive functions, and spans varying ranges of performances, POSET models allow for a flexible partial ordering of responses informing complex profiles of cognitive strengths and weaknesses. Theoretical statistical properties support this approach^30^, in that true latent profiles of mastery across cognitive functions will be correctly identified if the specifications are accurate, and enough task response data are observed.

As an illustration, the Category Fluency test asks participants to generate as many words as possible within a given semantic category, e.g., “vegetables.” Some functions required for the test include ideational fluency, word finding, rapid word retrieval, semantic memory, and executive functions (e.g., cognitive flexibility, effective search patterns). If an individual performs poorly on the Category Fluency test but performed well on another test, e.g., Boston Naming, which involves visual confrontational naming, poor performance on Category Fluency might then be attributed in part to poor executive functions (e.g., cognitive flexibility, effective search patterns), rather than a primary language deficit such as word-finding difficulty. Of course, variability in response behavior and measurement error must be considered when assessing the strength of this evidence. This inferential logic can be formalized within a statistical Bayesian framework. Importantly, this systematic approach can be extended to more complex scenarios, in which there are multiple functions involved within and across measures. Thus, POSET modeling integrates performance across measures to classify cognitive performance profiles, mimicking the logic of a neuropsychologist’s interpretation of confounded data.

The primary objective of this study is to provide a proof-of-concept demonstrating POSET modeling to characterize cognitive dysfunction in PD using the Parkinson Progression Markers Initiative (PPMI) data. As a secondary objective, we aim to evaluate the utility of POSET-derived scores in delineating cognitive trajectories and predicting future cognitive decline in patients who are cognitively normal at baseline.

## Results

The study cohort included 264 cognitively normal patients (67% male; mean age 61.16 years). At Year 3, 56 patients (21.2%) were classified as having cognitive dysfunction (51 with MCI, 5 with dementia), while 208 remained cognitively normal. The summary statistics (median and IQR) of the five CPS (Attention, Visuospatial Judgement, Executive Functioning, Working Memory, and Episodic Memory) across four visits (baseline, Years 1-3) for the 264 patients, stratified by presence or absence of cognitive dysfunction at Year 3, are provided as Table 3.

### Group Differences

Longitudinal trajectories of median CPS revealed distinct patterns between groups (Figure 1). Patients who remained cognitively normal at Year 3 (n = 208) demonstrated stable or slightly improved performance across all CPS over time. In contrast, patients who developed cognitive dysfunction by Year 3 (n = 56) exhibited markedly lower median CPS values that were evident as early as baseline despite being classified as cognitively normal. Mann-Whitney U tests confirmed significant differences between the groups. Patients who developed cognitive dysfunction by Year 3 had significantly lower CPS (p < 0.05) across all five CPS at Years 1-3. At baseline, group differences were significant for all CPS except Episodic Memory (p = 0.068); however, Episodic Memory CPS diverged significantly by Year 1 (eTables 2-5).

**Figure 1:**
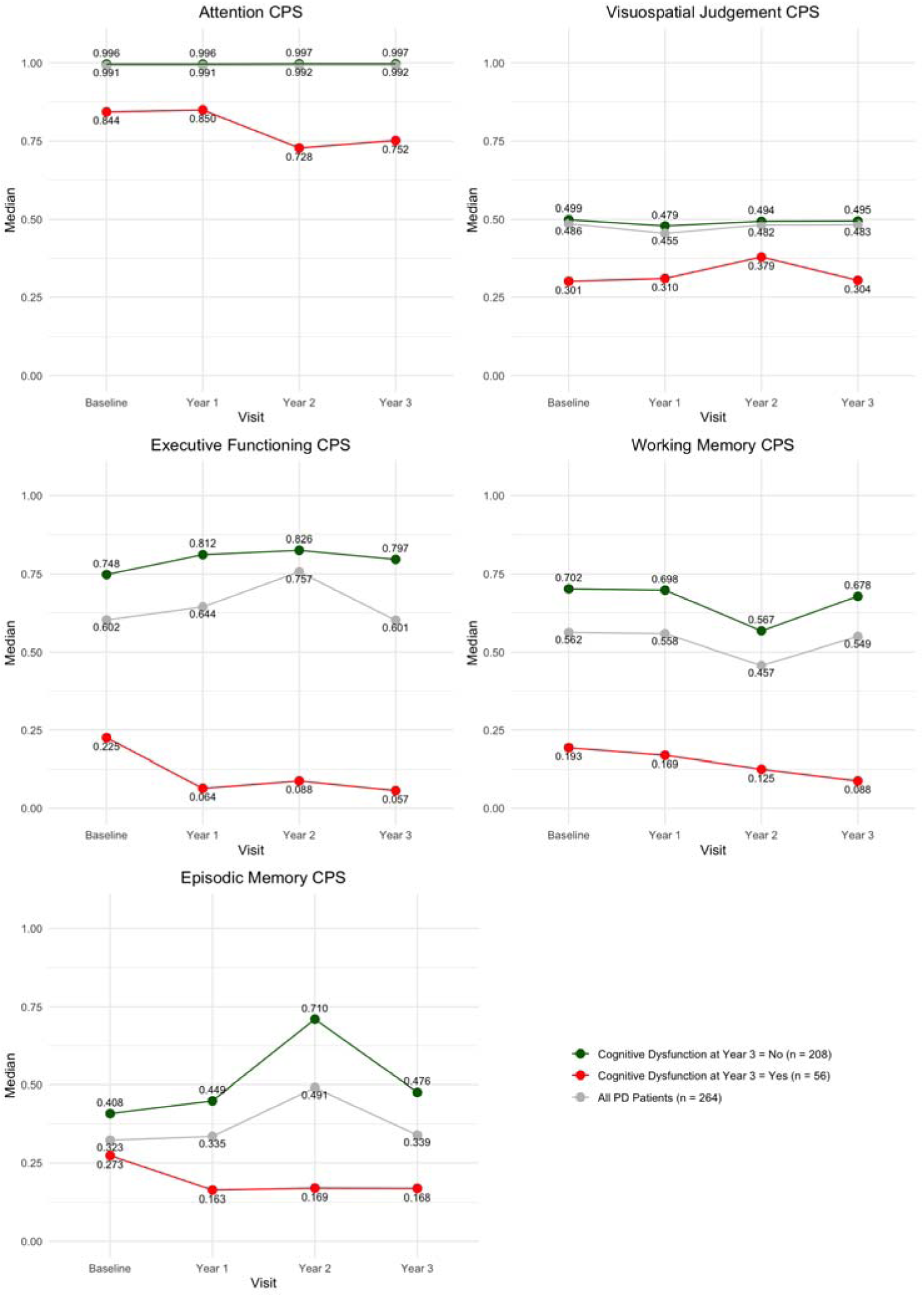
CPS Median Trajectories. Longitudinal trajectories of the median of the five Cognitive Performance Scores (CPS) across four visits, stratified by presence or absence of Cognitive Dysfunction at Year 3, with the full cohort included for reference.

### Prediction of Cognitive Dysfunction at Year 3

In the multivariable logistic regression model, lower baseline scores in Attention CPS (p = 0.049) and Visuospatial Judgement CPS (p = 0.030) were significant independent predictors of cognitive dysfunction at Year 3, with lower scores associated with higher odds of developing cognitive dysfunction by Year 3. Among the covariates, age at baseline was also a significant predictor (p = 0.011), while gender and education were not. The ROC analysis yielded an area under the curve (AUC) of 0.782 (95% confidence interval: 0.721, 0.843). Using an optimal probability cutoff of 0.40 to balance sensitivity and specificity, the model achieved 35.7% sensitivity (20/56), and 91.3% specificity (190/208). The p-value for Hosmer-Lemeshow test was > 0.05, suggesting adequate model fit.

## Discussion

This study demonstrates the feasibility of using POSET modeling to characterize the heterogeneity and trajectories of cognitive dysfunction in PD. By moving beyond binary classification and accounting for the polyfactorial nature of neuropsychological tests, we characterize distinct cognitive trajectories that precede clinical diagnosis.

A key strength of the POSET approach is its ability to reveal data-driven relationships between cognitive tests, scores, and operations that may be obscured by conventional clinical characterizations. For instance, in our model, the HVLT Recognition Discrimination score mapped to Executive Functioning. While clinically often viewed as a memory metric, the POSET model fit suggested that in this PD cohort, performance on this task was mathematically consistent with executive load, perhaps reflecting the cognitive demand of source monitoring or decision-making strategies. This highlights that POSET does not rely on expert consensus alone but uses the data structure to define how tests function in a specific cohort.

Among other analytical approaches reported in literature for predicting future cognitive dysfunction in PD, factor analysis has been the most widely used technique^31–34^. Factor analysis involves dimensionality reduction to identify subscales that are comprised of linear combinations of weighted cognitive test scores. Potential limitations include limited interpretability of subscales derived from factor loadings and imperfect reproducibility of loading patterns, particularly because different rotation choices applied to the same dataset can yield different solutions. When compared with factor analysis, which typically assumes a small number of continuous latent factors, POSET modeling characterizes the heterogeneity of performance as distinct cognitive profiles (partial ordered sets) and may provide additional insights into how neuropsychological tests invoke different cognitive domains. More generally, subscale approaches may not fully account for the polyfactorial nature of cognitive tests: poor performance on a polyfactorial measure can depress multiple subscale scores, even when the primary deficit reflects impairment in a function not well represented by a given subscale. Hence, subscale scores can be misleading. In contrast, POSET models explicitly represent polyfactorial test-domain relationships, and favorable statistical convergence properties^35^ support efficient and accurate identification of interpretable cognitive function mastery profiles.

Furthermore, the POSET models explicitly link cognitive functions and measures to provide a means for data-analytic validation of these links.^30^ Of note, other studies have also attempted to capture the richness of neuropsychological data into a single score for studying cognitive dysfunction in PD. Specifically, a recent study created a composite score by generating regression-based *z* scores for six cognitive tests for healthy controls and then averaging these *z* scores for patients with PD.^36^ In comparison, our Bayesian POSET models-based approach generates individual probabilities of mastery for specific functions that incorporate all associated tasks, recognizing the polyfactorial nature of tasks as opposed to enforcing a one-to-one association between a task and a domain. Importantly, the POSET models here are developed using patients with PD, so that differences and high performance are relative to those with PD, which may enhance the sensitivity to detect differences within PD. By contrast, combining modeling with healthy controls may make it more difficult to detect nuanced differences among patients with PD.

Our multivariate logistic regression analysis suggests that lower baseline Attention and Visuospatial Judgement CPS are independently associated with increased risk of cognitive dysfunction at Year 3, even after accounting for baseline demographic variables. These findings are consistent with prior work showing that early deficits in attention and visuospatial functioning may herald PD-MCI and PD-D^37^, and may also have implications for the synucleinopathy-based models of PD cognitive progression. For example, the significance of low baseline scores in the Visuospatial Judgement CPS in predicting conversion to cognitive dysfunction at Year 3 may be interpreted as consistent with modern neuropathological models and the proposed Dual Syndrome Hypothesis of PD^38^. Notably, the association of “prodromal” baseline loss of visuospatial performance despite overall clinician assessment of “normal cognition” with progression to dementia is congruent with caudal-to-rostral progression of Lewy body pathology^39–41^. Specifically, deficits in visuospatial performance align with the progression of Lewy body pathology into the parietal and temporal cortices and are consistent with the posterior-cortical syndrome that is more progressive and is the dementia-associated cognitive phenotype in PD^42^. This is complemented by the observation that other baseline CPS (e.g., Executive Functioning), while lower in converters, did not independently predict conversion, which is consistent with a more common and stable fronto-striatal cognitive phenotype predominated by executive dysfunction but less associated with progression to dementia^38^.

### Limitations

The exclusion of patients with baseline MCI or dementia was necessary to study incident decline, but may limit generalizability to the broader PD population. Future research should apply POSET modeling to cohorts encompassing the full spectrum of cognitive function in PD to characterize trajectories across different baseline cognitive states. Additionally, while using model-fit parameters to drive the mapping of neuropsychological tests to domains has inherent benefits, it consequently diverges from traditional neuropsychological categorizations in some instances (e.g., the exclusion of Immediate Recall from the Episodic Memory CPS). These mappings are specific to this dataset and cohort and thus should be interpreted as models and cohort-specific rather than definitive nosological claims.

Another potential limitation of this study stems from using the PPMI investigator-defined cognitive diagnosis (normal cognition, MCI, dementia) as the outcome, because the site investigator diagnostic decision, while fundamentally a clinical diagnosis, may have been informed by the neuropsychological tests used in the POSET models. This potential overlap could precipitate something of a circular approach wherein the neuropsychological tests used to map and generate the CPS may have also contributed to the ultimate decision of whether a participant had cognitive dysfunction. Ultimately, the site investigator’s diagnosis is foremost clinical, and while neuropsychological measures may have been incorporated into the medical decision making, investigator diagnosis remains a representative, closest-available gold standard of a holistic clinical assessment within PPMI.

Additionally, while the predictive model demonstrated good discriminative ability (AUC = 0.782) and achieved high specificity (91.3%), its sensitivity was low (35.7%). Clinically, the high specificity indicates that the model is effective at identifying patients who are unlikely to develop cognitive dysfunction within three years. Conversely, the low sensitivity suggests that the model missed a substantial proportion of patients who later developed cognitive dysfunction. Therefore, while these CPS provide useful predictive information, they are not sufficient alone for clinical prediction and should be integrated with other biomarkers and clinical risk factors to improve sensitivity for identifying at-risk individuals.

Other limitations include the lack of comprehensive assessment of cognitive dysfunction in the PPMI, a relatively short neuropsychological battery, and missing longitudinal data that may introduce sampling bias. To overcome some of these limitations, we are currently implementing and evaluating the POSET approach in a multi-center study, titled “Cognitive Functions Impairment as a Novel Paradigm for Delineating Cognitive Dysfunction in Parkinson’s Disease (PD-CFI)”^43^. Specifically, we are leveraging the data from the Pacific Northwest Udall Center of Excellence clinical consortium, designed specifically for studying cognitive dysfunction in PD^44^, along with consensus-based adjudication of cognitive dysfunction using the PD-MCI^2^ and PD-D^5^ criteria during consensus meetings among movement disorders neurologists and neuropsychologists, complemented by an informatics-based approach using a novel clinical decision support platform for PD (CDS-PD)^45^.

### Conclusion

POSET modeling offers a rigorous, mathematical framework for handling the complexity of neuropsychological data in PD. By generating continuous probability scores that reflect the underlying mastery of cognitive domains, this approach captures heterogeneity often lost in summary scores. Our proof-of-concept analyses using data from the PPMI study cohort demonstrate the feasibility of applying the POSET model-based approach to modeling the heterogeneity and trajectories of cognitive dysfunction in PD. This approach allows for the generation of individualized Cognitive Performance Scores (CPS) that capture subtle and specific differences in cognitive profiles (Figure 2). Notably, the distinct trajectories observed in Figure 1, particularly the early divergence of CPS medians between groups, underscore the sensitivity of this approach to early and differential cognitive changes. While this study was not designed to directly compare the POSET performance with conventional diagnostic criteria or other subtyping models, our findings in the PPMI cohort demonstrate the potential utility of this approach. Applying these models to larger and more comprehensive datasets may confirm these preliminary findings and help elucidate previously unknown factors in early PD that predict specific cognitive deficits later in the disease course.

**Figure 2:**
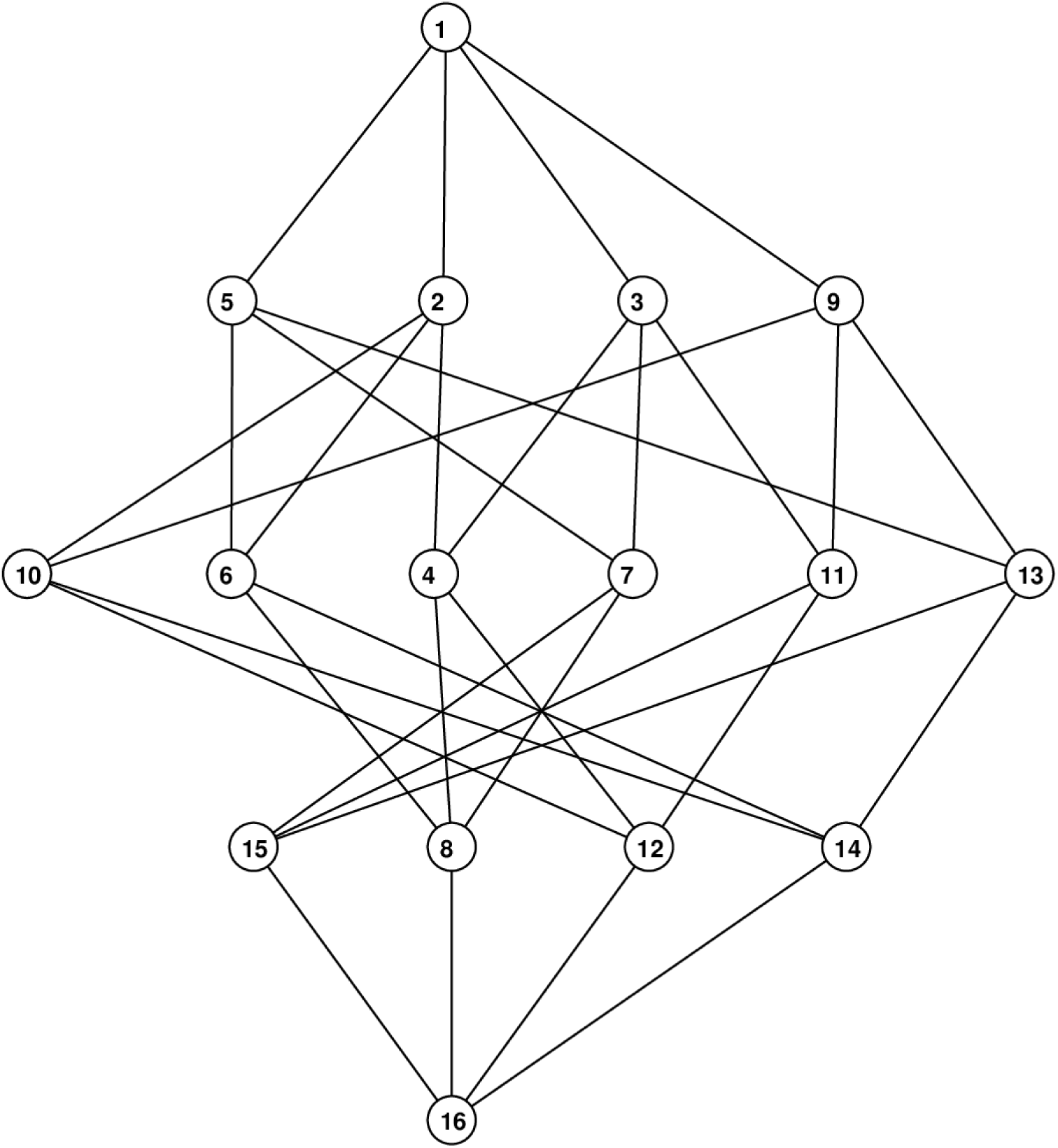
CPS POSET Model. Hasse diagram POSET model for cognitive functioning of subjects in PPMI data with individual numbers representing “States” from State 16 (lowest) to State 1 (highest).

## Methods

### Study Data and Patient Selection

We used data from 418 PD patients in the PPMI database as of June 2023, and categorized patients as having cognitive dysfunction if they were labeled as having either MCI or dementia by the site investigator at Year 3. We then selected 264 patients after excluding those with cognitive dysfunction at baseline or with any missing neuropsychological data from baseline to Year 3. Specifically, we only used cognitively normal patients at baseline to establish a clear starting point for observing the emergence of cognitive dysfunction later, allowing us to evaluate the predictive validity of baseline scores in an initially cognitively normal cohort and minimizing confounding from pre-existing impairment.

### POSET Score Mapping

We selected nine neuropsychological scores from five neuropsychological tests available in the PPMI dataset and used these to define five cognitive operations, representing the latent cognitive processes invoked by neuropsychological tests (Table 2): 1) Attention (the ability to attend to the demands of the cognitive battery), 2) Visuospatial Judgement (the ability to accurately perceive and interact with visuospatial information), 3) Executive Functioning (higher level cognitive skills that help guide behavior, such as planning, ideational fluency, problem-solving, and abstract thinking), 4) Working Memory (the ability to mentally manipulate information held in short-term memory) and 5) Episodic Memory (memory for recent or past events that involves encoding, storage, and retrieval of information).

**Table 2:**
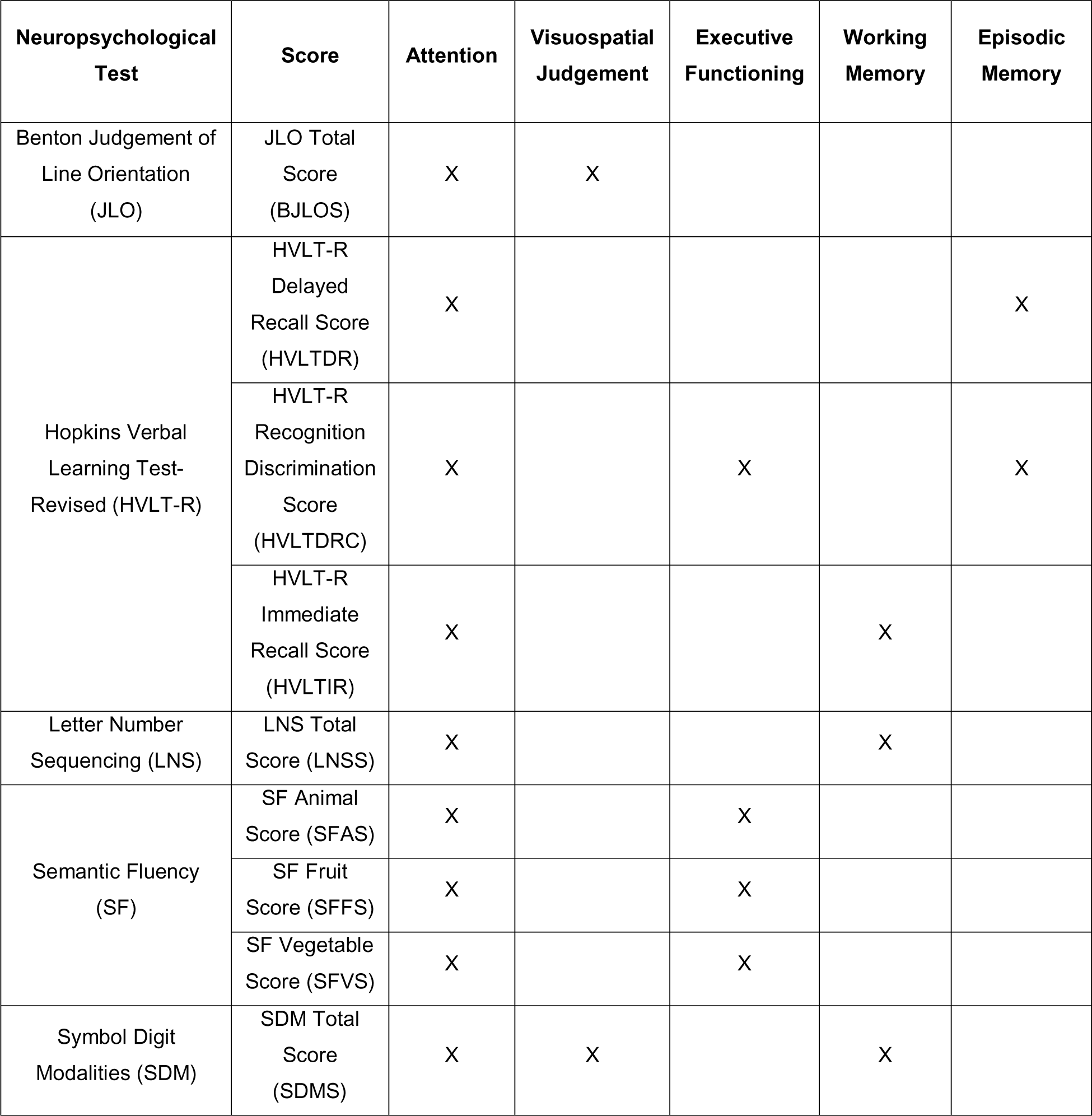
PPMI Neuropsychological Tests and Scores and their Mapping to Cognitive Operation. Five neuropsychological tests and corresponding nine scores available in the PPMI data, and their mapping to the cognitive operations.

**Table 3:** Descriptive Statistics for CPS Stratified by Cognitive Dysfunction. Median and IQR for the five CPS stratified by Cognitive Dysfunction (CD) and for all patients across four visits from baseline to Year 3.

| Cognitive Performance Score (CPS) |  | Median (Interquartile Range) |  |  |  |
| --- | --- | --- | --- | --- | --- |
|  |  | Baseline | Year 1 | Year 2 | Year 3 |
| Attention | CD = no | 0.996 (0.070) | 0.996 (0.101) | 0.997 (0.107) | 0.997 (0.093) |
|  | CD = yes | 0.844 (0.341) | 0.850 (0.449) | 0.728 (0.444) | 0.752 (0.460) |
|  | All Patients | 0.991 (0.142) | 0.991 (0.155) | 0.992 (0.211) | 0.992 (0.173) |
| Visuospatial Judgement | CD = no | 0.499 (0.497) | 0.479 (0.507) | 0.494 (0.493) | 0.495 (0.499) |
|  | CD = yes | 0.301 (0.210) | 0.310 (0.210) | 0.379 (0.208) | 0.304 (0.209) |
|  | All Patients | 0.486 (0.501) | 0.455 (0.494) | 0.482 (0.481) | 0.483 (0.497) |
| Executive Functioning | CD = no | 0.748 (0.889) | 0.812 (0.775) | 0.826 (0.845) | 0.797 (0.802) |
|  | CD = yes | 0.225 (0.746) | 0.064 (0.760) | 0.088 (0.887) | 0.057 (0.436) |
|  | All Patients | 0.602 (0.914) | 0.644 (0.898) | 0.757 (0.913) | 0.601 (0.906) |
| Working Memory | CD = no | 0.702 (0.683) | 0.698 (0.723) | 0.567 (0.757) | 0.678 (0.745) |
|  | CD = yes | 0.193 (0.653) | 0.169 (0.504) | 0.125 (0.313) | 0.088 (0.497) |
|  | All Patients | 0.562 (0.747) | 0.558 (0.758) | 0.457 (0.763) | 0.549 (0.815) |
| Episodic Memory | CD = no | 0.408 (0.804) | 0.449 (0.788) | 0.710 (0.711) | 0.476 (0.808) |
|  | CD = yes | 0.273 (0.495) | 0.163 (0.271) | 0.169 (0.344) | 0.168 (0.164) |
|  | All Patients | 0.323 (0.780) | 0.335 (0.766) | 0.491 (0.801) | 0.339 (0.801) |

The mapping of the neuropsychological scores to the cognitive operations was established through a data-driven process. While initial groupings were informed by standard neuropsychological concepts, the final mappings were determined by assessing the consistency of specifications to response data using POSET model-fit statistics^30^. This approach accounts for the specific behavior of tests within this dataset, sometimes resulting in mappings that differ from standard clinical taxonomy to optimize model fit. For example, while the Hopkins Verbal Learning Test-Revised (HVLT-R) Recognition Discrimination score is traditionally associated with Episodic Memory, analysis of statistical distributions (eFigures 1-3) and model fit in this cohort supported additional mapping to Executive Functioning, reflecting the possible executive demands of discrimination tasks in this population. Similarly, HVLT-R Immediate Recall trials were mapped to Attention and Working Memory driven by the model’s identification of attentional load.

### POSET Modeling

For each of the five cognitive operations, we calculated corresponding Cognitive Performance Scores (CPS) as POSET model derived posterior probabilities (ranging from 0 to 1) that estimates a participant’s mastery of a specific cognitive operation based on their performance across all mapped measures. A lower CPS indicates a lower probability of high-level functioning, whereas higher CPS values indicate greater probability of high level functioning.

### Outcome Definition

The main outcome measure was the presence of cognitive dysfunction at Year 3 (yes or no), defined as a diagnosis of either MCI or dementia by the site investigator.

### Standard Protocol Approvals, Registrations, and Patient Consents

This study used deidentified data from the PPMI, which is overseen and approved by the WCG Institutional Review Board (IRB) and for which written informed patient consent was obtained from all participants.

### Statistical Analysis

Statistical analyses were performed using IBM SPSS Statistics for Macintosh, Version 29.0.1.1. For reporting the characteristics of the raw neuropsychological scores, we used mean and standard deviation (eTable 1). Given the non-normal distribution of CPS values (eFigures 4-8), we utilized the Mann-Whitney U test to compare median CPS values between groups: results are summarized as median and interquartile range (IQR).

We also conducted a multivariate logistic regression to examine whether the five CPS at baseline would be predictive of cognitive dysfunction at Year 3, with baseline age, gender, and education as covariates. Model fit statistics were calculated, and classification performance was evaluated. To further evaluate the discriminative ability of the model for Year 3 cognitive dysfunction, we conducted a Receiver Operating Characteristic (ROC) analysis to determine area under the curve (AUC), sensitivity, and specificity. Statistical significance was set at p < 0.05.

## Data Availability Statement

Data used in the preparation of this article were obtained on [2023-06-28] from the Parkinson’s Progression Markers Initiative (PPMI) database (www.ppmi-info.org/access-data-specimens/download-data), RRID:SCR_006431. For up-to-date information on the study, visit www.ppmi-info.org.

The dataset used in this publication will be made available by request from any qualified investigator.

## Supporting information

eTables and eFigures

## Acknowledgements

This study was funded by the project titled “Cognitive Functions Impairment as a Novel Paradigm for Delineating Cognitive Dysfunction in Parkinson’s Disease (PD-CFI)”, funded by the Investigator Initiative Research Award (IIRA) of the Parkinson Research Program (PRP) of the Congressionally Directed Medical Research Program (CDMRP) of the United States (US) Department of Defense (DoD). Award # HT94252310222

We thank the PPMI – a public-private partnership – which is funded by the Michael J. Fox Foundation for Parkinson’s Research, and funding partners. Multiple authors are employees of The VA Parkinson’s Disease Research, Education, and Clinical Care Centers (PADRECC).

## Author’s Roles

C. Zweber: Conceptualization, Data Curation, Formal Analysis, Investigation, Validation, Visualization, Writing – Original Draft, Writing – Review & Editing

JJ: Methodology, Validation, Writing – Review & Editing

C. Zabetian: Writing – Review & Editing

RM: Methodology, Validation, Writing – Review & Editing

VI: Writing – Review & Editing

AH: Writing – Review & Editing

SS: Writing – Review & Editing

BC: Methodology, Validation, Writing – Review & Editing

AR: Writing – Methodology, Validation, Review & Editing

CT: Conceptualization, Formal Analysis, Investigation, Visualization, Writing – Review & Editing

DKG: Conceptualization, Funding Acquisition, Investigation, Project Administration, Supervision, Methodology, Writing – Original Draft, Writing – Review & Editing

