## Supplementary material for "Modeling the Heterogeneity and Trajectories of Cognitive Dysfunction in Parkinson’s Disease Using Partially Ordered Set Models": eTables and eFigures

**eTable 1: Descriptive Statistics of Original Nine PPMI Neuropsychological Test Measures**

| **Neuropsychological Test Score** | **Mean (standard deviation)** | | | |
| --- | --- | --- | --- | --- |
|  | **Baseline (n=418)** | **Year 1 (n=365)** | **Year 2 (n=360)** | **Year 3 (n=325)** |
| BJLOS | 12.76 (2.13) | 12.41 (2.37) | 12.78 (2.28) | 12.66 (2.25) |
| HVLTDR | 8.35 (2.52) | 8.15 (2.79) | 8.16 (2.98) | 8.36 (3.01) |
| HVLTDRC | 13.63 (2.63) | 10.68 (2.54) | 12.64 (2.45) | 11.92 (2.35) |
| HVLTIR | 24.44 (4.98) | 23.78 (5.33) | 23.59 (5.50) | 24.81 (6.03) |
| LNSS | 10.59 (2.66) | 10.44 (2.62) | 10.26 (2.82) | 10.25 (3.04) |
| SFAS | 20.95 (5.35) | 20.96 (5.41) | 20.96 (5.78) | 20.86 (5.52) |
| SFFS | 13.50 (4.08) | 13.47 (4.20) | 13.71 (4.67) | 13.62 (4.38) |
| SFVS | 14.22 (4.52) | 14.26 (4.29) | 14.25 (4.72) | 13.80 (4.18) |
| SDMS | 41.20 (9.73) | 40.93 (10.10) | 39.83 (11.17) | 39.89 (11.68) |

Caption: BJLOS = Benton Judgement of Line Orientation Total Score; HVLTDR = Hopkins Verbal Learning Test Revised - Delayed Recall Score; HVLTDRC = Hopkins Verbal Learning Test Revised - Recognition Discrimination Score; HVLTIR = Hopkins Verbal Learning Test Revised - Immediate Recall Score; LNSS = Letter Number Sequencing Total Score; SFAS = Semantic Fluency Animal Score; SFFS = Semantic Fluency Fruit Score; SFVS = Semantic Fluency Vegetable Score; SDMS = Symbol Digit Modalities Total Score.

**eTable 2: Baseline CPS Stratified by Cognitive Dysfunction with Mann-Whitney Testing**

| **Cognitive Performance Score (CPS)** | **Baseline (Median, IQR)** | | **Mann-Whitney** | |
| --- | --- | --- | --- | --- |
|  | **CD = no**  **(n = 208)** | **CD = yes**  **(n = 56)** | **U** | **p-value** |
| Attention | 0.996 (0.070) | 0.844 (0.341) | 3102.5 | **< 0.001** |
| Visuospatial Judgement | 0.499 (0.497) | 0.301 (0.210) | 3690.5 | **< 0.001** |
| Executive Functioning | 0.748 (0.889) | 0.225 (0.746) | 4039.0 | **< 0.001** |
| Working Memory | 0.702 (0.683) | 0.193 (0.653) | 3445.5 | **< 0.001** |
| Episodic Memory | 0.408 (0.804) | 0.273 (0.495) | 4897.0 | 0.068 |

Caption: Medians, interquartile ranges, and distribution significance for the baseline CPS, split by presence of Cognitive Dysfunction (CD) at Year 3. Significant (p < 0.05) bolded.

**eTable 3: Year 1 CPS Stratified by Cognitive Dysfunction with Mann-Whitney Testing**

| **Cognitive Performance Score (CPS)** | **Year 1 (Median, IQR)** | | **Mann-Whitney** | |
| --- | --- | --- | --- | --- |
|  | **CD = no**  **(n = 208)** | **CD = yes**  **(n = 56)** | **U** | **p-value** |
| Attention | 0.996 (0.070) | 0.850 (0.449) | 3235.5 | **< 0.001** |
| Visuospatial Judgement | 0.479 (0.507) | 0.310 (0.210) | 4705.0 | **0.027** |
| Executive Functioning | 0.812 (0.775) | 0.064 (0.760) | 3440.5 | **< 0.001** |
| Working Memory | 0.698 (0.723) | 0.169 (0.504) | 3006.0 | **< 0.001** |
| Episodic Memory | 0.449 (0.788) | 0.163 (0.271) | 4164.5 | **0.001** |

Caption: Medians, interquartile ranges, and distribution significance for the year 1 CPS, split by presence of Cognitive Dysfunction (CD) at Year 3. Significant (p < 0.05) bolded.

**eTable 4: Year 2 CPS Stratified by Cognitive Dysfunction with Mann-Whitney Testing**

| **Cognitive Performance Score (CPS)** | **Year 2 (Median, IQR)** | | **Mann-Whitney** | |
| --- | --- | --- | --- | --- |
|  | **CD = no**  **(n = 208)** | **CD = yes**  **(n = 56)** | **U** | **p-value** |
| Attention | 0.997 (0.107) | 0.728 (0.444) | 3085.0 | **< 0.001** |
| Visuospatial Judgement | 0.494 (0.493) | 0.379 (0.208) | 4431.5 | **0.006** |
| Executive Functioning | 0.826 (0.845) | 0.088 (0.887) | 3651.0 | **< 0.001** |
| Working Memory | 0.567 (0.757) | 0.125 (0.313) | 3167.5 | **< 0.001** |
| Episodic Memory | 0.710 (0.711) | 0.169 (0.344) | 3429.0 | **< 0.001** |

Caption: Medians, interquartile ranges, and distribution significance for the year 2 CPS, split by presence of Cognitive Dysfunction (CD) at Year 3. Significant (p < 0.05) bolded.

**eTable 5: Year 3 CPS Stratified by Cognitive Dysfunction with Mann-Whitney Testing**

| **Cognitive Performance Score (CPS)** | **Year 3 (Median, IQR)** | | **Mann-Whitney** | |
| --- | --- | --- | --- | --- |
|  | **CD = no**  **(n = 208)** | **CD = yes**  **(n = 56)** | **U** | **p-value** |
| Attention | 0.997 (0.093) | 0.752 (0.460) | 2331.0 | **< 0.001** |
| Visuospatial Judgement | 0.495 (0.499) | 0.304 (0.209) | 4213.0 | **0.001** |
| Executive Functioning | 0.797 (0.802) | 0.057 (0.436) | 2737.0 | **< 0.001** |
| Working Memory | 0.678 (0.745) | 0.088 (0.497) | 2815.5 | **< 0.001** |
| Episodic Memory | 0.476 (0.808) | 0.168 (0.164) | 3889.0 | **< 0.001** |

Caption: Medians, interquartile ranges, and distribution significance for the Year 3 CPS, split by presence of Cognitive Dysfunction (CD) at Year 3. Significant (p < 0.05) bolded.

**eFigures**

**eFigure 1: HVLT-R Delayed Recall and Recognition Discrimination Overall Performance**

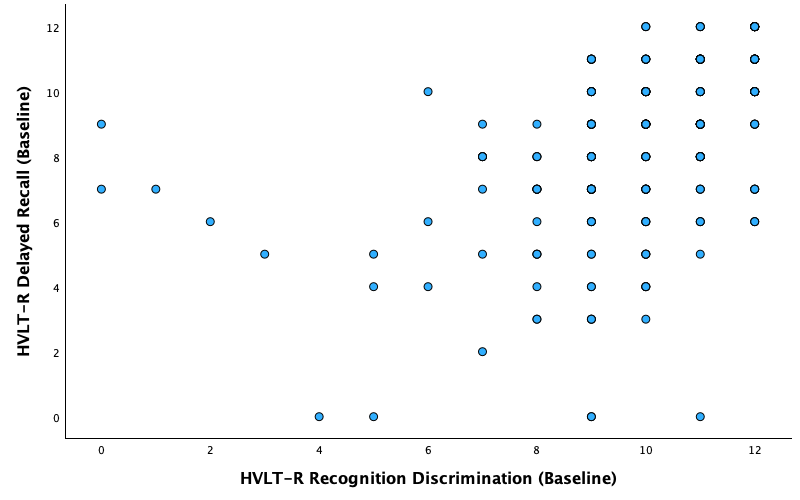

Caption: Scatter Plot of Baseline HVLT-R Delayed Recall and HVLT-R Recognition Discrimination Scores at Baseline for All Patients.

**eFigure 2: HVLT-R Delayed Recall and Recognition Discrimination Without Cognitive Dysfunction Performance**

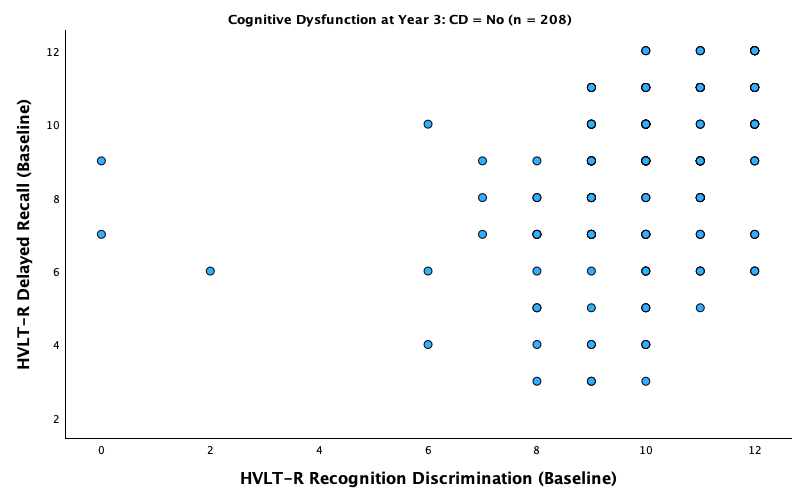

Caption: Scatter plot of HVLT-R Delayed Recall and HVLT-R Recognition Discrimination Scores at baseline for patients without Cognitive Dysfunction.

**eFigure 3: HVLT-R Delayed Recall and Recognition Discrimination With Cognitive Dysfunction Performance**

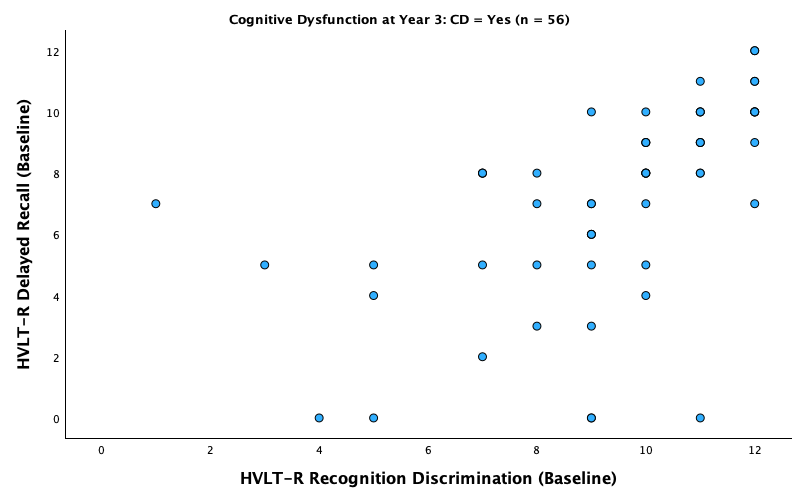

Caption: Scatter plot of HVLT-R Delayed Recall and HVLT-R Recognition Discrimination Scores at baseline for patients with Cognitive Dysfunction.

**eFigure 4: Distribution of Baseline Attention CPS**

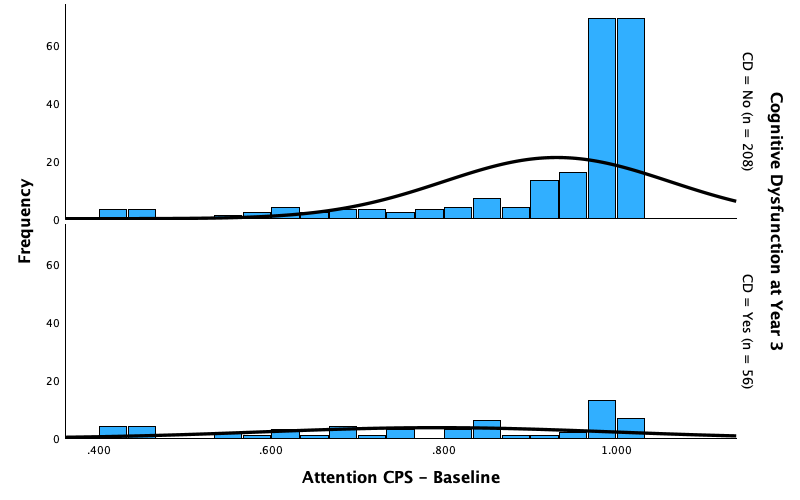

Caption: Distribution of baseline Attention CPS, stratified by Cognitive Dysfunction at Year 3. Normal curve shown in black.

**eFigure 5: Distribution of Baseline Visuospatial Judgement CPS**

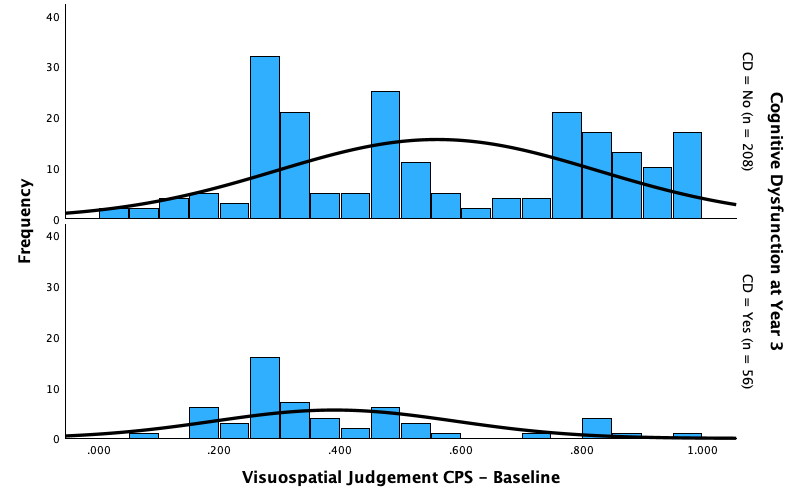

Caption: Distribution of baseline Visuospatial Judgement CPS, stratified by Cognitive Dysfunction at Year 3. Normal curve shown in black.

**eFigure 6: Distribution of Baseline Executive Functioning CPS**

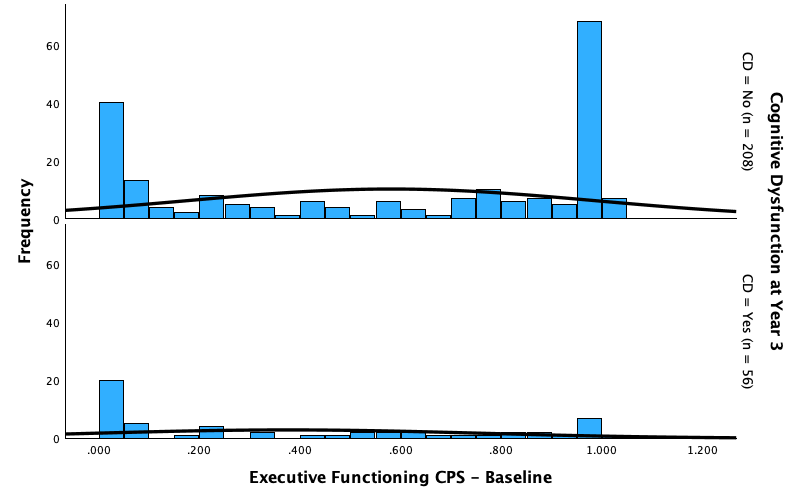

Caption: Distribution of baseline Executive Functioning CPS, stratified by Cognitive Dysfunction at Year 3. Normal curve shown in black.

**eFigure 7: Distribution of Baseline Working Memory CPS**

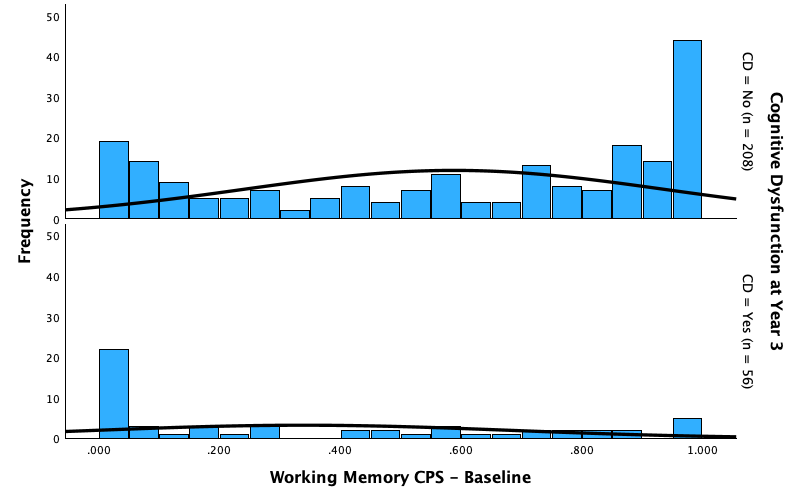

Caption: Distribution of baseline Working Memory CPS, stratified by Cognitive Dysfunction at Year 3. Normal curve shown in black.

**eFigure 8: Distribution of Baseline Episodic Memory CPS**

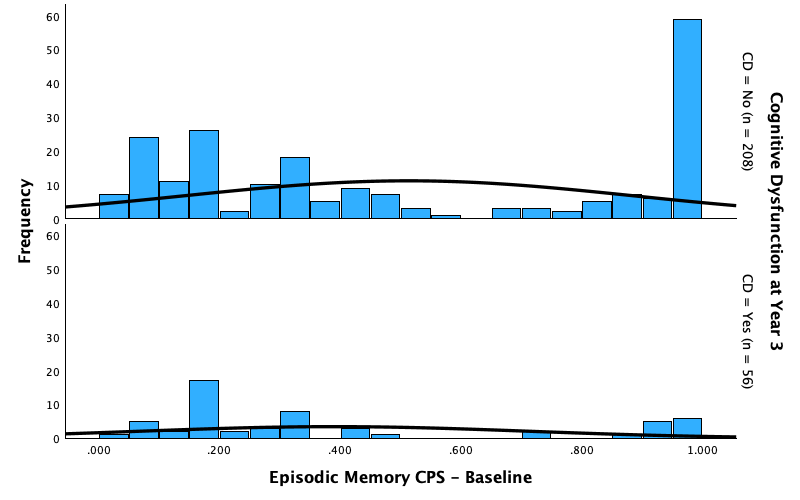

Caption: Distribution of baseline Episodic Memory CPS, stratified by Cognitive Dysfunction at Year 3. Normal curve shown in black.
